# Inflammation in children with chronic kidney disease linked to gut dysbiosis and metabolite imbalance

**DOI:** 10.1101/2022.01.21.22269663

**Authors:** Johannes Holle, Hendrik Bartolomaeus, Ulrike Löber, Felix Behrens, Theda U. P. Bartolomaeus, Harithaa Anandakumar, Moritz I. Wimmer, Dai Long Vu, Mathias Kuhring, Andras Maifeld, Sabrina Geisberger, Stefan Kempa, Philip Bufler, Uwe Querfeld, Stefanie Kitschke, Denise Engler, Leonard D. Kuhrt, Oliver Drechsel, Kai-Uwe Eckardt, Sofia K. Forslund, Andrea Thürmer, Victoria McParland, Jennifer A. Kirwan, Nicola Wilck, Dominik Müller

## Abstract

Chronic kidney disease (CKD) is characterized by a sustained pro-inflammatory response. The underlying mechanisms are incompletely understood, but may be linked to gut dysbiosis. Dysbiosis has been described in adults with CKD; however, comorbidities limit CKD-specific conclusions. We analyzed the fecal microbiome, metabolites and immune phenotypes in children at three different CKD stages (G3-G4, G5 (hemodialysis), after kidney transplantation) and healthy controls. Serum TNF-α and sCD14 were stage-dependently elevated, indicating inflammation and gut barrier dysfunction. We observed microbiome alterations in CKD, including a diminished production of short-chain fatty acids. Bacterial tryptophan metabolites were increased in CKD. CKD serum activated the aryl hydrocarbon receptor and stimulated TNF-α production by monocytes, corresponding to a shift towards intermediate/non-classical monocytes. Unsupervised T cell analysis revealed pro-inflammatory shifts in MAIT and Treg cells. Thus, gut barrier dysfunction and microbial metabolites exacerbate inflammation and may therefore contribute to the increased cardiovascular burden in CKD.

## Introduction

Despite ongoing efforts to improve the treatment of patients with chronic kidney disease (CKD), they still suffer from high morbidity and mortality, primarily due to cardiovascular diseases (CVD)^1^. Besides known risk factors such as arterial hypertension and proteinuria^1^, recent studies suggest a crucial role for microbially produced metabolites in promoting inflammation^2,3^ and, as a consequence, progression of CKD and CVD^1,3,4^.

There is longstanding evidence showing that an imbalance in the bacterial community residing in the gut with changes in its functional composition, termed dysbiosis, is common in adult patients with CKD^5,6^. Beyond the influence of CKD, a variety of other factors, such influence of diet and drugs, are suspected to contribute to dysbiosis in CKD^6^. These changes are paralleled by an altered bacterial metabolism of nutrients and a systemic accumulation of uremic toxins of bacterial origin, such as indoxyl sulfate (IxS)^7^. It is conceivable that dysbiosis and metabolite dysbalance aggravate systemic inflammation, which could provide a potential mechanism for the high rate of premature cardiovascular events^3^.

Recently, we demonstrated a positive association between IxS and early cardiovascular disease in children with CKD^8^. In contrast to adults with CKD, children are less affected by risk factors such as diabetes, obesity, and metabolic syndrome, but mainly suffer from congenital anomalies of the kidney and urinary tract (CAKUT)^9^. Thus, a pediatric cohort offers the unique opportunity to analyze the impact of CKD on microbiota-host interaction more specifically.

Our cohort includes pediatric CKD patients, those on hemodialysis (HD), patients after kidney transplantation (KT) and age-matched healthy controls (HC). We show, for the first time a CKD specific dataset of gut microbiome composition, altered microbial metabolism of nutrients, and the corresponding impact on inflammation and immune cell dysregulation in children suffering from CKD. We connect the bacterial metabolite dysbalance to a pro-inflammatory immune cell signature, emphasizing the importance of the microbiota for chronic inflammation in CKD. The fact that these dysbiosis-driven immunological changes are already detectable in children with CKD highlights the potential of microbiota-targeted therapies to improve prognosis of CKD patients across all ages.

## Patients and methods

### Study population

In this cross-sectional study, we recruited patients from the Department of Pediatric Gastroenterology, Nephrology, and Metabolic Diseases at Charité University hospital in Berlin, Germany, between February 2018 and June 2018. Written informed consent was obtained from all participants and/or their parents prior to study entry. The study was approved by the local Ethical Review Board (EA2/162/17). All procedures performed were in accordance with the ethical standards of the institutional and national research committees and the 1964 Helsinki declaration and its later amendments or comparable standards.

Patients (age 3-18 years) were enrolled in the following groups:

- CKD group: CKD stage G3-G4, estimated GFR (eGFR) 15-60 ml/min*1.73m^2^
- HD group: CKD stage G5D, patients on maintenance hemodialysis, enrolled earliest four weeks after initiation of HD
- KT group: patients after successful KT, earliest four weeks after KT, without a history of rejection or chronic graft failure, eGFR > 60ml/min*1.73m^2^
- HC group: normal kidney function, treated at the hospital for reasons other than kidney disease

We excluded patients with a body weight below 15kg, acute or chronic inflammatory diseases, fever, diabetes, chronic liver disease, inflammatory bowel disease, or other gastrointestinal disorders (constipation, diarrhea, short bowel syndrome). Patients with antibiotic prophylaxis or treatment within the four weeks prior to recruitment were excluded.

### Statistical analysis

#### Microbiome and metabolome analysis

Alpha diversities of microbial communities (Shannon diversity as computed by RTK, defined at the OTU level) were compared between groups using Kruskal-Wallis (KW) test. Beta diversity was assessed using Euclidean (metabolome) and Canberra (microbiome) dissimilarity index between samples computed using vegan package v2.5-7. Principal Coordinates Analysis (PCoA) was performed using the vegan package v2.5-7 (employing Euclidean and Canberra distance metrics as above). PERMANOVA was performed using the adonis function from the vegan package v2.5-7. We estimate differential abundance on phylum and genus level between groups using the package DESeq2 v1.30.1. The DESeq2 pipeline uses negative binomial distribution models to test for differential abundance between testing conditions. We ran the pipeline with normalized counts under default settings. *P* values were adjusted according to Benjamini-Hochberg (BH) false discovery rates (FDR) correction. A q-value of <0.1 was considered statistical significant.

For each pair of patient groups, features from the TRP analysis were compared using two-sided Mann-Whitney-U (MWU) tests with effect sizes calculated as Cliff’s delta metric per the R ordom package v3.1. Effect sizes were taken as Spearman’s rho. All significance estimates were adjusted for multiple tests using BH-FDR correction. To assess the effect of patient groups on tryptophan pathway metabolites (concentrations), multifactor ANOVAs were calculated per metabolite to account for multiple groups as well as potential confounders (including age, sex, ethnical background, underlying disease category, BMI, and eGFR).

The co-abundance network of host, microbiome and metabolome features was calculated from the dataset as a whole by assessing pairwise Spearman correlations and adjusted for multiple testing using BH-FDR correction as implemented in the R psych package v1.9.12. Edges for which absolute rho > 0.3 and *Q* < 0.1 were visualized using the iGraph R package. Correlations were separately also assessed stratifying for group effects using the R coin package v1.3.1.

#### Flow cytometry

For all FlowSOM clusters we computed log2 fold changes (fc). Cluster log2fc were visualized using ggplot2 package v3.3.5. For all subpopulations of Treg and MAIT cells, we calculated log2fc and assessed statistical significance using two-sided MWU-tests between HD and HC with BH-FDR correction. A q-value of <0.1 was considered statistically significant. Data was visualized using the EnhancedVolcano package v4.1.2.

#### Boxplots

Analysis and graphical representation was performed in GraphPad Prism 9.3.1 (GraphPad Software, CA, USA). Boxplots depict median and interquartile range with whiskers from min to max. Overlaid data points represent individual patients or measurements. Normality was assessed using Q-Q-plots and Shapiro-Wilk test. For more than two groups we performed one-way ANOVA with Tukey post-hoc test or KW test with Dunn’s post-hoc test, as appropriate. For two group comparisons, we performed two-sided Student’s t-test or two-sided MWU test, as appropriate.

For all analysis *P* value of <0.05 (unadjusted or adjusted, as appropriate) and a *Q* value of <0.1 was considered statistically significant.

## Results

### Childhood CKD marked by arterial hypertension, inflammation and leaky gut

Ten healthy individuals (HC) and 38 patients were enrolled in the study (Supplementary Figure 1). Baseline characteristics are given in Table 1. Participant mean age was 10.6 ± 3.8 years with 21 females and 27 males. CAKUT was the most prevalent underlying disease category. Patients treated with HD had a median time on dialysis of six months (range 3 – 29 months) with a median residual diuresis of 50 ml per day (range 0 – 1800 ml per day) and a mean Kt/Vof 1.56 ± 0.27 (SD), indicating adequacy of HD treatment. Patients after KT had a stable graft function with a mean eGFR of 78.8 ± 19.4 ml/min*1.73m^2^. The median time from transplantation was 48 months (range 10 – 125 months).

**Table 1:** Patients baseline characteristics. Patients (n = 48) were grouped into four categories (CKD = chronic kidney disease, HD = hemodialysis, KT = kidney transplantation, HC = healthy controls). Data is shown as mean ± standard deviation or percentage (%) as appropriate.

| Patients (n) | HC<br>10 | CKD G3-G4<br>12 | HD<br>11 | KT<br>15 |
| --- | --- | --- | --- | --- |
| Age (years) | 12.1 ± 4.2 | 8.3 ± 2.3 | 13.6 ± 3.2 | 9.1 ± 2.9 |
| Female | 7 (70%) | 5 (42%) | 3 (27%) | 6 (40%) |
| Diagnosis |  |  |  |  |
| CAKUT | N/A | 5 (42%) | 5 (45%) | 8 (53%) |
| Tubulointerstitial | N/A | 2 (17%) | 1 (9%) | 3 (20%) |
| Glomerulopathy | N/A | 4 (33%) | 4 (36%) | 3 (20%) |
| Post-AKI | N/A | 1 (8%) | 1 (9%) | 1 (7%) |
| Healthy | 10 (100%) | N/A | N/A | N/A |
| Caucasian | 9 (90%) | 12 (100%) | 3 (27%) | 11 (73%) |
| Weight (percentile) | 61 ± 23 | 38 ± 21 | 8 ± 7 | 52 ± 28 |
| BMI (percentile) | 57 ± 21 | 35 ± 19 | 31 ± 19 | 59 ± 25 |
| eGFR (ml/min/1.73 m<sup>2</sup>) | 100.2 ± 29.3 | 29.6 ± 14.6 | 6.6 ± 1.1 | 78.6 ± 19.4 |
| Urea (mg/dl) | 30 ± 7 | 112 ± 67 | 147 ± 39 | 36 ± 14 |
| Uric acid (mg/dl) | 4.8 ± 1.3 | 7.6 ± 2.2 | 6.7 ± 1.2 | 5.4 ± 1.4 |
| Phosphate (mmol/l) | 1.3 ± 0.1 | 1.6 ± 0.3 | 2.0 ± 0.5 | 1.5 ± 0.3 |
| Albumin (g/l) | 47.7 ± 5.3 | 43.8 ± 13.0 | 38.0 ± 4.8 | 40.0 ± 3.4 |
| CrP (mg/l) | 2.6 ± 5.9 | 0.7 ± 1.1 | 3.6 ± 6.9 | 2.0 ± 1.8 |
| PTH (pmol/l) | N/A | 13.2 ± 8.5 | 38.5 ± 38.4 | 7.7 ± 3.8 |
| Triglycerides (mg/dl) | N/A | 166 ± 75 | 186 ± 54 | 112 ± 57 |
| Antihypertensive treatment | 0 (0%) | 10 (83%) | 10 (91%) | 8 (53%) |
| Antihypertensive drugs (n) | 0 | 1.8 ± 1.5 | 2.4 ± 1.9 | 0.8 ± 0.9 |
| Arterial Hypertension<br>(BP > 95<sup>th</sup> Percentile) | N/A | 2 (17%) | 6 (55%) | 5 (33%) |

Abbreviations: CAKUT = congenital anomalies of the kidney and urinary tract; Post-AKI = patients with chronic kidney disease after acute kidney injury; BP = blood pressure; N/A = not applicable.

Both the CKD group (CKD stage G3-G4) and the HD group (CKD stage G5D) were hypertensive (83% and 91%, respectively) and received antihypertensive treatment (1.8 ± 1.5 antihypertensive drugs in the CKD group and 2.4 ± 1.9 drugs in the HD group, Figure 1a, Table 1). Six HD patients (55%) exhibited hypertensive blood pressures at the time of study enrolment despite treatment (Table 1). In the HD and CKD groups serum levels of the pro-inflammatory cytokine TNF-α were stage-dependently increased and significantly higher than in the HC group (Figure 1b), while differences in the levels of other pro- and anti-inflammatory cytokines did not reach significance (Supplementary Table 2). Notably, TNF-α was also significantly elevated in patients after KT compared to HC (Figure 1b). We next investigated serum levels of the tight junction protein zonulin-1 (Zo-1) and the lipopolysaccharide (LPS) binding protein soluble CD14 (sCD14), which were elevated in CKD and HD compared to control and KT (Figure 1c), indicating stage-dependent intestinal barrier dysfunction and CKD-associated endotoxemia. Thus, in the absence of classical cardiovascular risk factors other than hypertension, CKD in children is characterized by elevated serum markers of inflammation and leaky gut.

**Figure 1:**
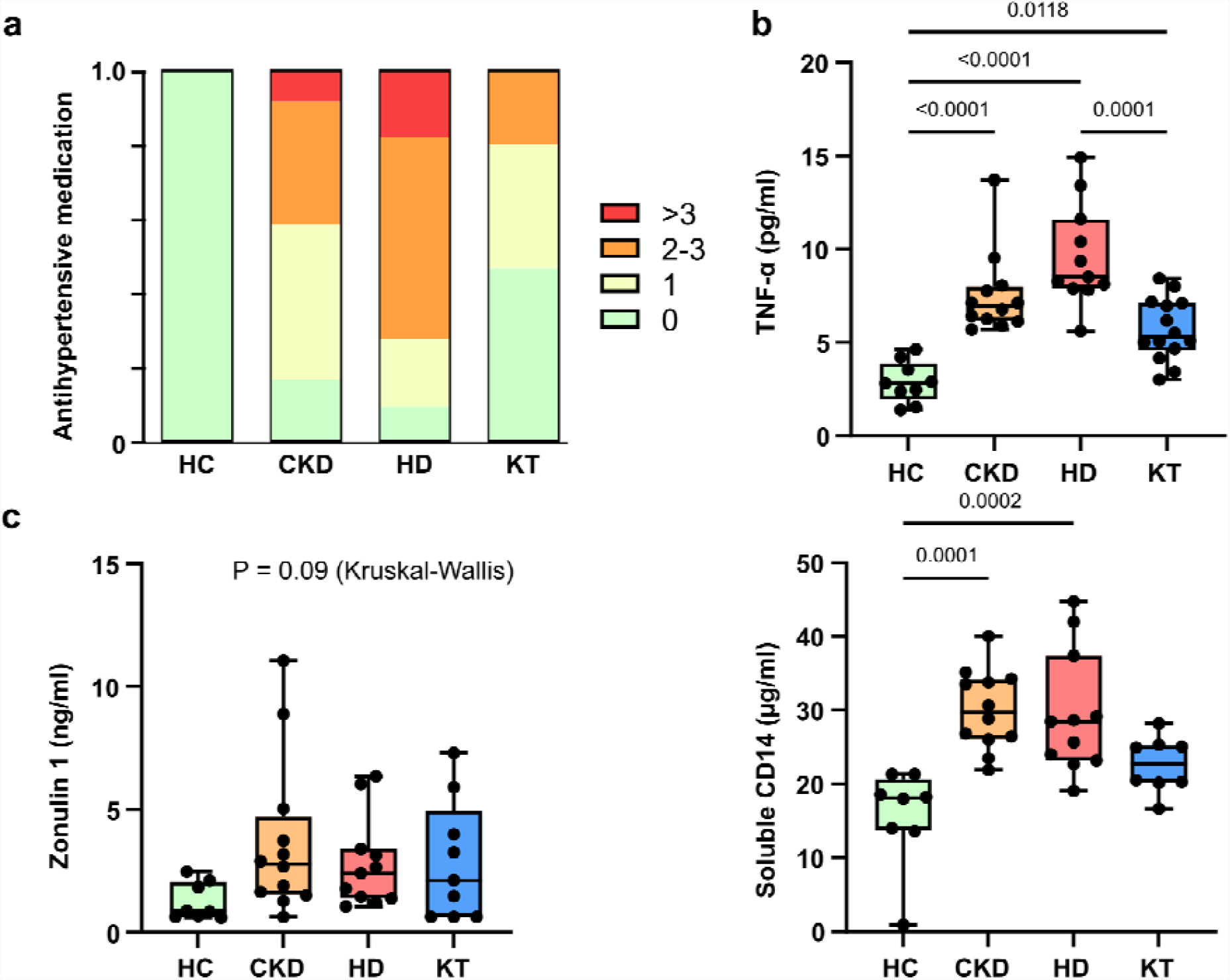
Arterial hypertension and systemic inflammation are linked to impaired intestinal barrier function in pediatric chronic kidney disease. The number of antihypertensive drugs per individual (a, n= 48 patients) is shown in patients with chronic kidney disease (CKD G3-4), hemodialysis (HD), after kidney transplantation (KT) and healthy controls (HC). Plasma TNF-α (b, n=46 patients) was analyzed by chemiluminescence immunoassay. Gut barrier function was assessed using Zonulin 1 and soluble CD14 (c, n=40 patients) ELISA measurements in plasma. Data is shown as a box (median and interquartile range) and whiskers (min-max) with overlaid dot plot. *P* values ≤ 0.05 are shown, as measured by ordinary one-way ANOVA or Kruskal-Wallis test followed by Tukey’s or Dunn’s post-hoc correction for multiple comparisons, as appropriate.

### Microbiome alterations in CKD

Since intestinal barrier dysfunction may be associated with dysbiosis, we next sought to analyze the taxonomic composition of the gut microbiome. We performed 16S amplicon sequencing in n=32 study participants with available fecal samples. We observed a high interindividual compositional variability at phylum level (Figure 2a). Microbiome richness and diversity, as measured by the Shannon diversity index, tended to be lower in CKD and HD patients compared to HC but this difference did not reach statistical significance (Figure 2b). However, analysis of beta diversity (Canberra distance) indicated significant differences in microbiome composition between groups, with CKD and HD groups separating most clearly from the HC (Figure 2c, p = 0.01). Analysis of bacterial composition on genus level revealed significant alterations predominantly in HD patients (Figure 2d). First, relative abundances of Firmicutes and Actinobacteria such as *Fusicatenibacter* (belonging to the family of Lachnospiraceae), *Subdoligranulum* (Ruminococcaceae), and *Bifidobacterium* (Bifidobacteriaceae) were significantly diminished in HD patients compared to HC. Second, we found an increase in relative abundance of Proteobacteria and Bacteroidetes, such as *Citrobacter* (Enterobacteriaceae), *Parasutterella* (Parasutterellaceae), and several genera of *Bacteroides* (Bacteroidaceae) in CKD and HD groups compared to HC and KT. Taken together, the taxonomic microbiome changes in CKD are stage-dependent, most pronounced in the HD group and less pronounced after KT. The abundance of proteolytic bacteria (such as *Citrobacter*) increases in CKD whereas saccharolytic bacteria (such as *Bifidobacterium*) decrease.

**Figure 2:**
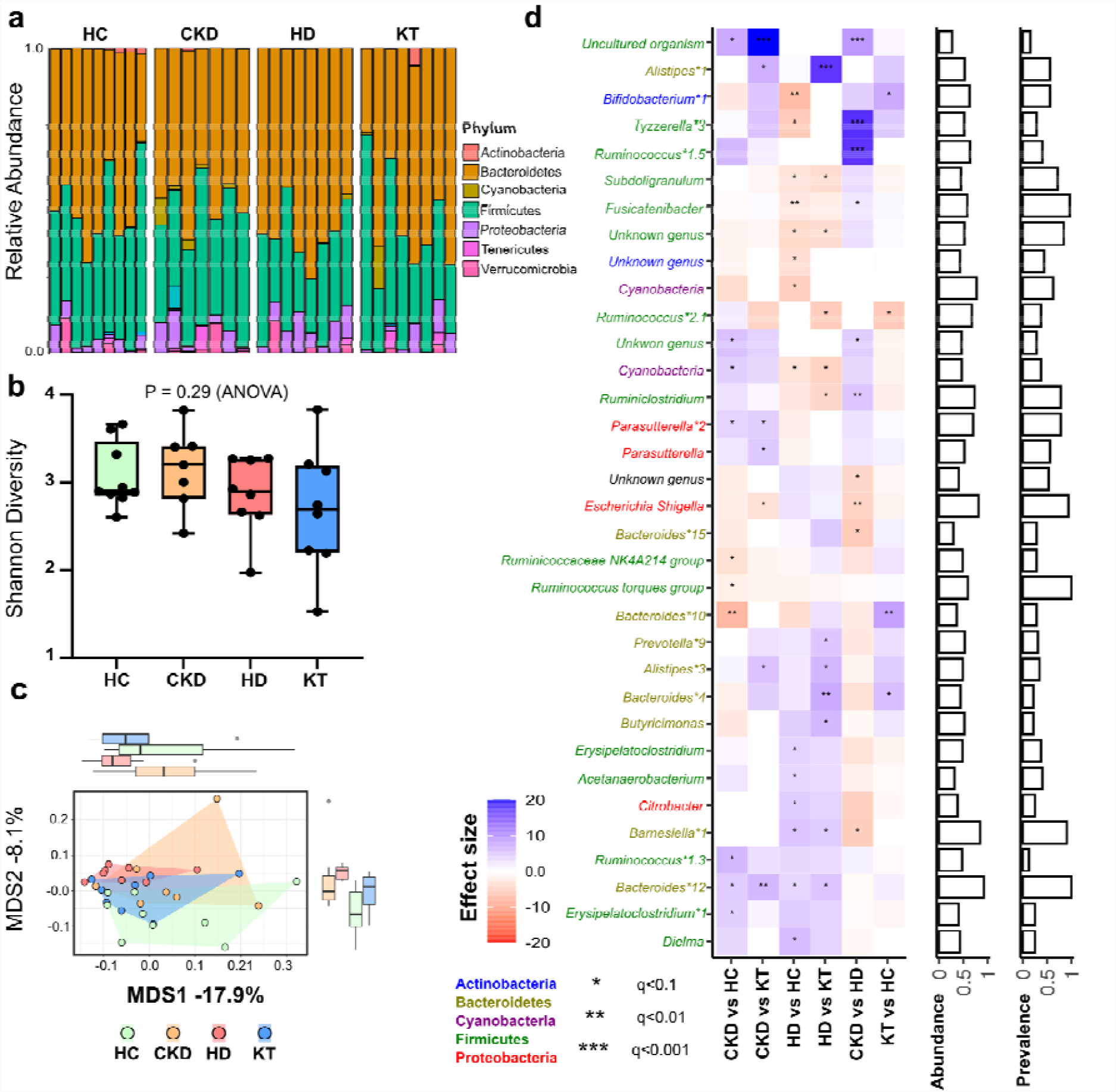
Characteristics of gut microbiota in a cohort of pediatric patients with chronic kidney disease compared to healthy controls. Analysis of gut microbiota from 16S rRNA sequencing in children (n = 32) with chronic kidney disease (CKD G3-4), patients with hemodialysis (HD), patients after kidney transplantation (KT) and healthy controls (HC). a) Relative abundance on phylum level of individuals according to their respective group. b) Alpha diversity as measured by Shannon diversity. Data is shown as a box (median and interquartile range) and whiskers (min-max) with overlaid dot plot. c) Beta diversity assessment by Principal Coordinate Analysis (PCoA) based on Canberra distance (p = 0.01 by PERMANOVA). d) Analyses of group differences on genus level are shown as a heatmap. Patient groups were tested against each other (pairwise). The heatmap shows significant changes in abundance using DESeq2 v1.30.1 package. Multiple groups of the same genus reported by Lotus due to lacking coverage in available phylogenetic databases are marked by numbers. Bar charts (right) show abundance and prevalence; abundance is calculated as log(genus count)/log(max(genus count)); prevalence for each genus is calculated across the whole data set. All significance estimates were adjusted for multiple tests using Benjamini-Hochberg FDR correction.

### CKD stage-dependent dysbalance of bacterial metabolites

To investigate the functional effects of the observed alterations in microbiome composition on host physiology, we focused a plasma metabolite analysis on tryptophan (TRP) metabolites and SCFA. Dietary TRP is a substrate for both cellular metabolism and bacterial proteolytic fermentation and the latter is a source of microbial produced uremic toxins, such as IxS. We found significant differences in the abundance of TRP metabolites in CKD and HD compared to the HC and KT groups (Figure 3a). CKD and HD patients showed a large-scale shift from TRP to its indole and kynurenine (KYN) metabolites, which was predominantly driven by an increase of IxS (Figure 3b). We found a significant decrease in plasma TRP concentrations in patients with CKD and HD, while IxS and 5 KYN metabolites (KYN, kynurenic acid (KA), 3-OH-kynurenine (3OH-KYN), anthranilic acid (AA), xanthurenic acid (XA)) were significantly elevated (Figure 3c). Interestingly, patients after KT had similar metabolite levels compared to HC with the exception of KYN and 3OH-KYN (Figure 3c). Individual values of TRP, IxS and, KA are shown across the different groups in Figure 3d-f. The activation of the cellular KYN pathway is also indicated by the ratio of KYN to TRP, which was significantly elevated in CKD and HD (Figure 3g). A multivariate ANOVA showed that age, sex, ethnic background, underlying disease category, and BMI did not confound metabolite levels. Only group and eGFR had an impact on several of the compounds (Supplementary Table 3), confirming their accumulation with declining eGFR. Because indole and KYN metabolites have been shown to activate the aryl hydrocarbon receptor (AhR), a transcription factor and potential mediator of microbiome-host interactions, we measured the AhR activating potential of serum from the HD and HC group *in vitro* using a cell-based reporter assay. Serum from HD patients induced a significantly higher AhR activity as compared to HC (Figure 3h). Similarly, IxS activates the AhR in a dose-dependent manner (Supplementary Figure 2).

**Figure 3:**
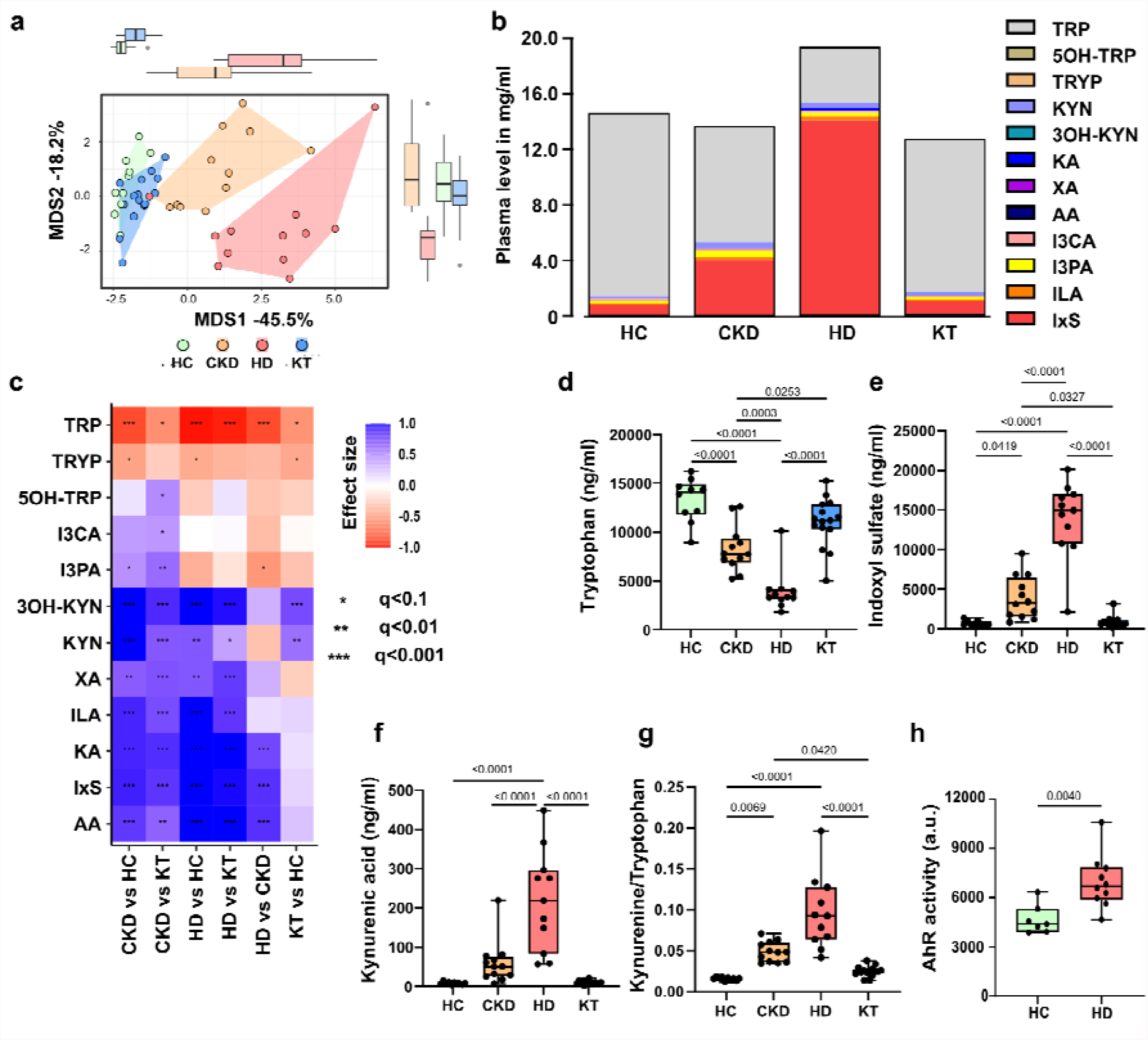
Stage-dependent activation of plasma tryptophan metabolism activates the Aryl-hydrocarbon receptor. Tryptophan (TRP) and its metabolites were measured in plasma of children at different stages of chronic kidney disease (CKD) compared to healthy controls (n = 48). a) Multivariate analysis (Principal Coordinate Analysis) of all measured metabolites discriminates between patients with CKD G3-4, patients with hemodialysis (HD), patients after kidney transplantation (KT) and healthy controls (HC). b) Cumulative load of TRP and its metabolites. c) Univariate analysis depicted as heatmap shows effect sizes (Cliff’s delta) for each pair of patient groups. Colours denote the effect directions (blue-positive and red-negative) and magnitudes (the darker the colour, the stronger the magnitude); asterisks represent the association significance. Statistical significance was assessed by Mann-Whitney U-test and Benjamini-Hochberg false discovery rate correction. Group differences of TRP (d), indoxyl sulfate (IxS, e) and kynurenin acid (KA, f) were further visualized in box plots. The Kynurenine/Tryptophan ratio (g) indicates the activity of tryptophan degradation to kynurenine metabolites. The activity of the Aryl-hydrocarbon receptor (AhR, h) was analyzed using a transfected reporter cell line after 48 hrs incubation with serum of HC (n=7) and HD (n=10) patients. *P* values ≤ 0.05 are shown, as measured by ordinary one-way ANOVA or Kruskal-Wallis test and adjusted by post-hoc Tukey’s or Dunn’s correction for multiple testing (d-g) or by t test (h). Data is shown as a box (median and interquartile range) and whiskers (min-max) with overlaid dot plot. Abbreviations: TRP= tryptophan, 5OH-TRP= 5-hydroxy-tryptophan, TRYP= tryptamin, KYN= kynurenine, 3OH-KYN= 3-hydroxy-kynurenine, KA= kynurenic acid, AA= anthranilic acid, XA= xanthurenic acid, IxS= indoxyl sulfate; I3CA= indole-3-carboxyaldehyde, I3PA= indole-3-propionic acid, ILA= indole lactate, AhR= aryl hydrocarbon receptor.

To further functionally validate our findings of decreased abundancies of saccharolytic microbes, we measured the abundance of a central enzyme for bacterial SCFA production by qPCR of fecal DNA using degenerate primers. The abundancy of the butyrate gene was lower in the DNA extracted from fecal samples of HD patients relative to the other patient groups (Supplementary Figure 3a). Next, we measured SCFA in serum. The SCFA propionate and isobutyrate were reduced in patients with HD compared to HC, while serum concentrations of butyrate did no differ significantly between groups (Supplementary Figure 3b). Thus, we were able to show on a functional level that the observed stage-dependent microbiome changes have an impact on the production and abundance of bacterial metabolites.

In order to comprehensively visualize the relationship of clinical, microbial and metabolomic parameters, we performed a correlation network analysis (Figure 4). Here, TNF-α correlated positively with sCD14, Proteobacteria, IxS, several KYN metabolites and biomarkers of kidney function (Figure 4a, positive correlations). Conversely, the SCFA propionate and isobutyrate correlated inversely with TNF-α, IxS, KYN metabolites and kidney function (Figure 4b, negative correlations). Firmicutes correlated positively with microbial diversity, butyrate and I3PA. These associations support our assumption of kidney function as a catalyst of gut bacteria-driven inflammation.

**Figure 4:**
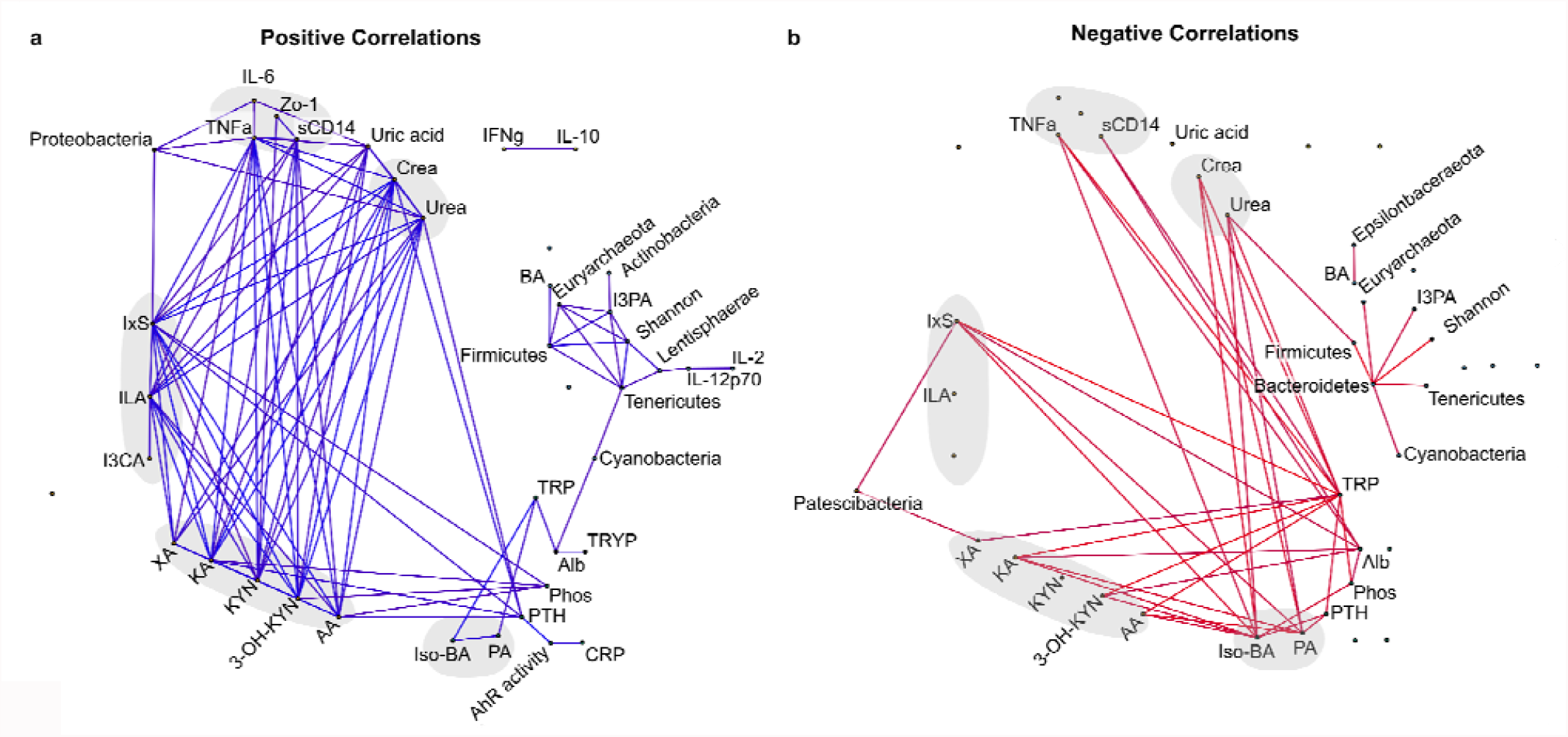
Correlation network of gut microbiome, clinical parameters, plasma tryptophan metabolites and cytokines. Laboratory parameters, tryptophan metabolites (n = 48), cytokines (n=46) and taxonomic data (n = 32) were associated using pairwise Spearman correlations and adjusted for multiple testing using the Benjamini-Hochberg FDR correction. Edges for which absolute rho > 0.3 and Q < 0.1 are visualized. For better visualization eGFR was removed as creatinine and urea convey similar information. a) Positive correlations. b) Negative correlations. Abbreviations: TRYP= tryptamin, TRP= tryptophan, KYN= kynurenine, KA= kynurenic acid, 3OH-KYN= 3-hydroxy-kynurenine, AA= anthranilic acid, XA= xanthurenic acid, PA= propionic acid, BA= butyric acid, Iso-BA= isobutyric acid, Zo-1= zonulin-1, sCD14= soluble CD14, IxS= indoxyl sulfate, ILA= indole lactate, I3CA= indole-3-carboxyaldehyde, I3PA= indole-3-propionic acid, CrP= C-reactive protein. Crea= creatinine, phos= phosphate, Alb= albumin.

### Monocyte subsets contribute to the pro-inflammatory phenotype in CKD

The AhR is expressed in various immune cells including myeloid cells and is known to modulate their function^10^. It is assumed that the accumulation of AhR ligands in CKD leads to immune cell activation. We therefore isolated monocytes from healthy donors and incubated them with serum from HD patients or HC. Monocytes incubated with HD serum showed a significantly higher TNF-α production (Figure 5a). Similarly, incubation of isolated monocytes with IxS dose-dependently increased TNF-α production which could be reversed by co-incubation with the synthetic AhR-antagonist CH-223191, highlighting the importance of AhR-mediated immune activation in CKD (Figure 5a).

**Figure 5:**
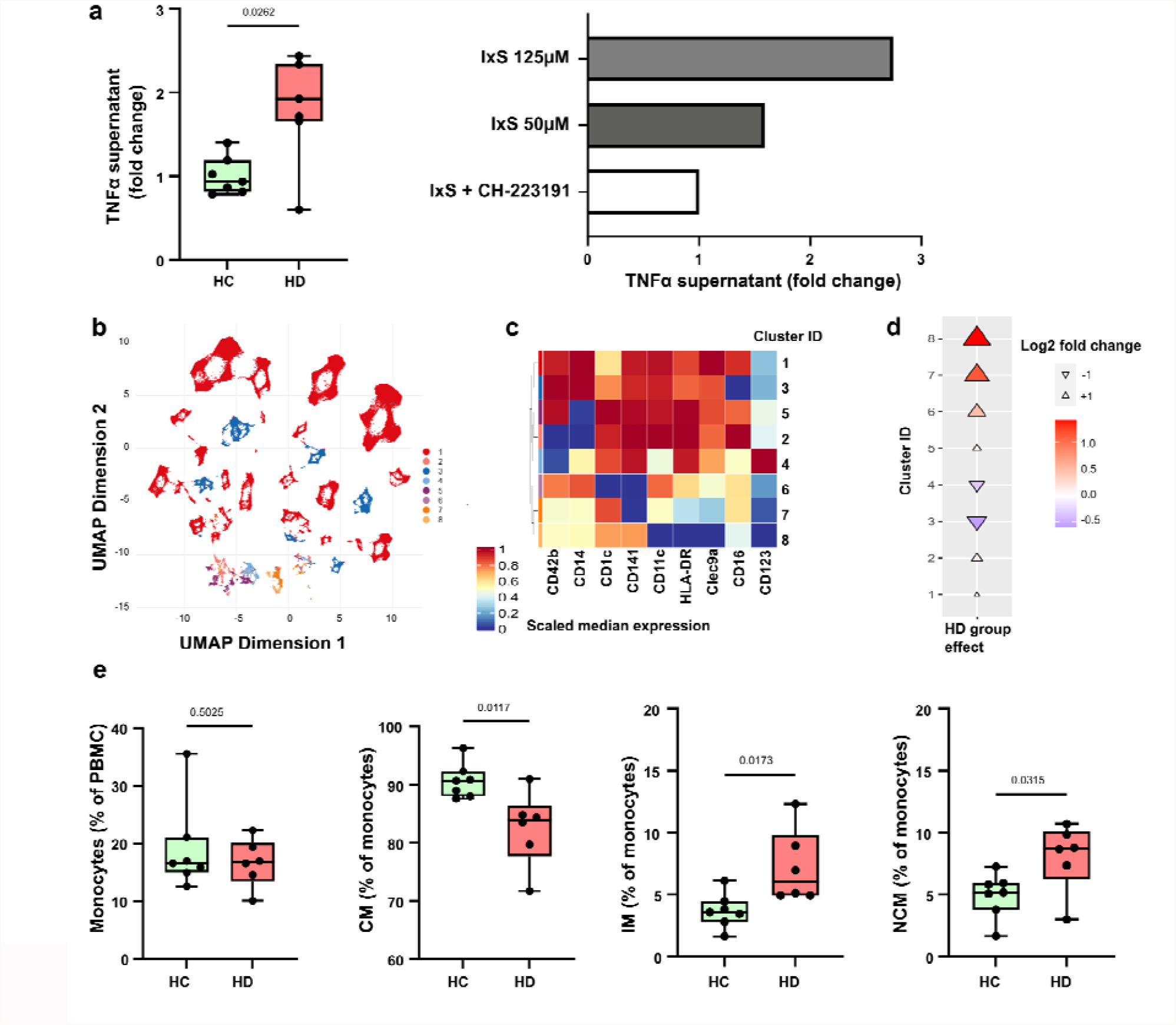
Monocyte subtypes promote inflammation in chronic kidney disease. Monocytes isolated from healthy donors were incubated with serum from hemodialysis patients (HD, n=7) and healthy controls (HC, n=7). Monocytes were incubated with indoxyl sulfate (IxS) in presence or absence of the AhR antagonist CH-223191. TNF-α was measured in the culture supernatant after 24 hrs incubation using ELISA (a). Peripheral blood mononuclear cells (PBMC) were isolated from HD (n=6) and HC (n=7) individuals for surface staining and multi-color flow cytometry was performed. Unsupervised clustering by FlowSOM revealed 8 different cell clusters (b) characterized by the differential expression of 9 surface marker describing myeloid and dendritic cells (c). Cuneiform plots depict the log2-fold changes for these clusters between HD and HC (indicated by color, size and directionality of the triangles, d). Classical hierarchical gating of total monocytes, classical monocytes (CD16+), non-classical (CD14+), and intermediate (CD14+CD16+) monocytes is shown in e). Abbreviations: AhR= aryl hydrocarbon receptor.

In order to get a broader overview of changes in relevant immune cell populations in CKD, we re-collected peripheral blood mononuclear cells (PBMC) from seven HC and six HD patients for immunophenotyping using flow cytometry. Unsupervised FlowSOM analysis^11^ of our monocyte and dendritic cell targeting flow panel (see Supplementary table 1, supplementary figure 4, Figure 5b), showed phenotypic alterations of monocytes (cluster 3, Figure 5c) being decreased (Figure 5d) and dendritic cell (cluster 7, Figure 5c) being increased (Figure 5d) in HD patients. Using classical hierarchical gating, we observed similar abundances of total monocytes (identified by HLA-DR, CD14, and CD16 as described in^12^), but a significant shift from classical (CD14+CD16-) towards non-classical (CD14-CD16+) and intermediate (CD14+CD16+) monocytes in HD (Figure 5e), the latter two being known for their potent production of TNF-α^13^. Hierarchical gating within the dendritic cell population revealed no significant differences of dendritic cell types (Supplementary Figure 5). Thus, HD patients are characterized by alterations within myeloid cells, showing a shift from classical to pro-inflammatory intermediate and non-classical monocytes.

### Pro-inflammatory T cell subsets in CKD

Since SCFA and TRP metabolites are known to modulate T cell differentiation and function, we expanded our flow cytometry to include analysis of T cells (Supplementary table 1, T surface panel, supplementary figure 4). We again performed FlowSOM analysis using 8 clusters (Figure 6a). Mucosal associated invariant T (MAIT) cells and a subpopulation of regulatory T cells (Treg) (Clusters 6 and 3, respectively, Figure 6b) exhibited the largest effects between HD and HC patients (Figure 6c). We confirmed this finding by classical hierarchical gating, with circulating MAIT cells (CD3+CD161+TCRVα7.2+) being decreased in HD (Supplementary Figure 6a). Next, we performed a more detailed analysis of MAIT cell subpopulations. After adjusting for multiple testing, we found a decrease of CD4-CD8-MAIT cells, while CD4+CD8- and CD4+CD8+ cells were enriched in HD (Figure 6d, f). Moreover, MAIT cells of HD patients displayed an effector memory phenotype (CD45RA-, CD62L-) and expressed more PD-1, indicative of increased activation. Importantly, upon *in vitro* restimulation, MAIT cells of HD patients secreted significantly more interleukin-17A (IL-17A) (Figure 6d, f).

**Figure 6:**
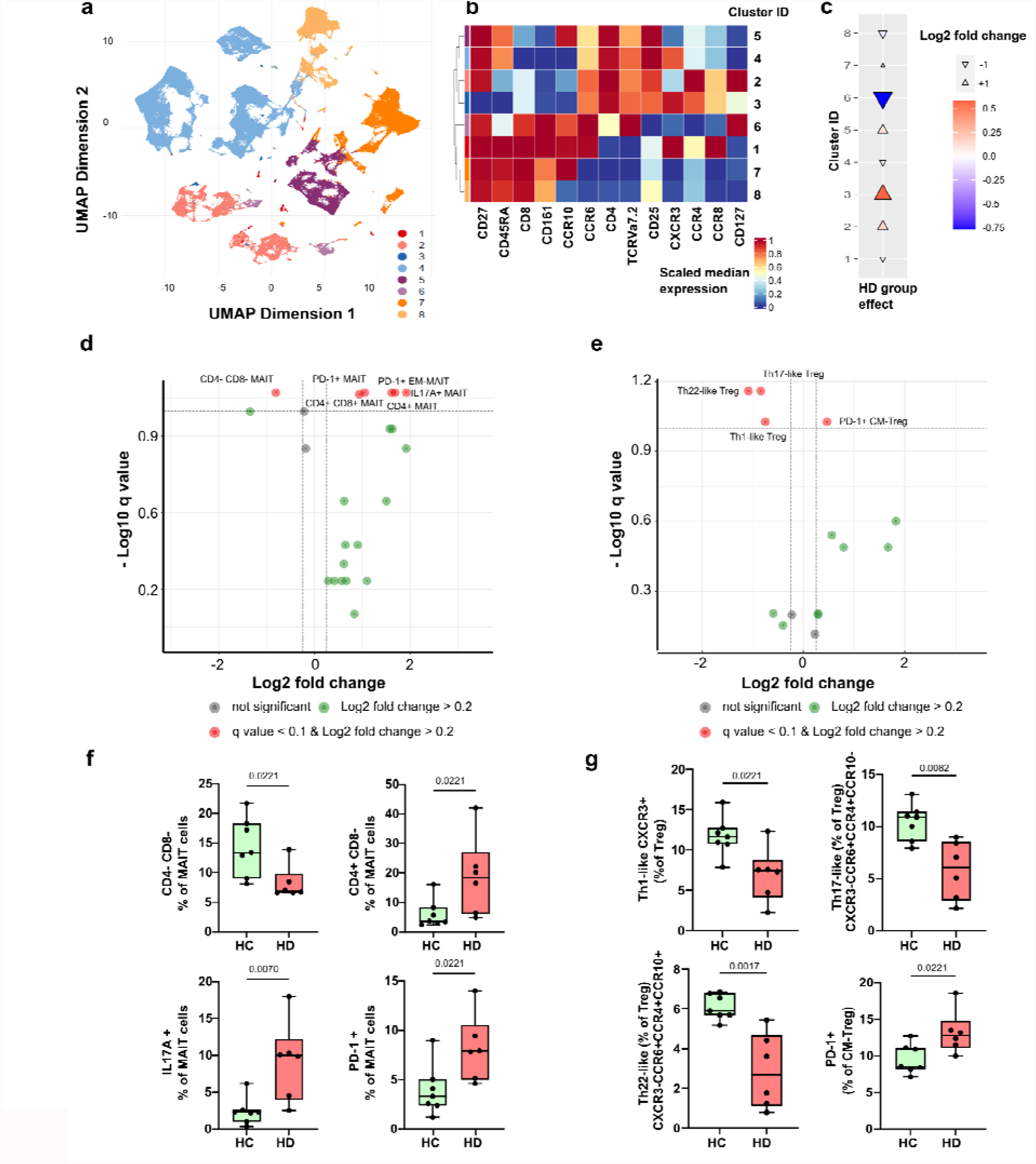
Pro-inflammatory T cell subtypes in chronic kidney disease. Unsupervised clustering of HD (n=6) and HC (n=7) individuals by FlowSOM revealed 8 T cell clusters (a) based on the differential expression of surface marker (b). c) Cuneiform plots showing the log2-fold changes for these clusters between HD and HC (indicated by color, size and directionality of the triangles). Volcano plot of MAIT (d) and Treg (e) subpopulations by hierarchical gating. y-axis indicates Q value by Mann-Whitney U-test and Benjamini-Hochberg false discovery rate correction, x-axis log2-fold change between HD and HC. Significantly altered subpopulations are depicted as box (median and interquartile range) and whiskers (min-max) with overlaid dot plots for MAIT and Treg in f) and g), respectively. For f and g, *P* values ≤ 0.05 are shown, as measured by t test or Mann-Whitney-U test as appropriate. Abbreviations: MAIT= mucosa-associated invariant T cells; Treg= regulatory T cells, HD= hemodialysis, HC= healthy controls.

In addition, we identified changes within a CD4+CD25+CD127-cell cluster (Cluster 3, Figure 6a-c), indicating alterations in Treg subpopulations. Total Treg proportions were not altered between HC and HD (Supplementary Figure 6b). However, since different subpopulations with partly different functions are known to exist within the total Treg population, we extended the Treg characterization (Supplementary table 1, T surface and T activation)^14^. Adjusted for multiple testing, several Treg subpopulations, characterized by activation markers and chemokine receptors, were significantly changed in HD patients (Figure 6e). In detail, Treg from HD patients with a central-memory phenotype (CD45RA-, CD62L+) showed a higher expression of PD-1 indicating activation state. Based on the expression of chemokine receptors, Th1-like (CXCR3+), Th17-like (CXCR3-CCR6+CCR4+CCR10-) and Th22-like (CXCR3-CCR6+CCR4+CCR10+) Treg were significantly diminished in HD patients compared to HC (Figure 6e, g). Taken together, we observed significant alterations in T cell subtypes, namely MAIT cells and Treg, which play an important role in mucosal immunity and inflammation.

## Discussion

Inflammation is a hallmark of CKD, detectable even at young age and particularly prognosis-determining. We show in children with CKD, that intestinal barrier dysfunction and dysbiosis are associated with a systemic bacterial metabolite imbalance that contributes to the pro-inflammatory phenotype of several immune cells including monocytes and T cells. We also provide novel insight into the microbiota-immune crosstalk mediated by TRP metabolites and SCFA in CKD. Thus, we demonstrate a stage-dependent aberration of the microbiome-immune axis, which is both a contributor to inflammation and a potential target for anti-inflammatory therapeutic strategies.

The young cohort enrolled for this study exhibits a markedly increased risk for cardiovascular disease, indicated by a high prevalence of arterial hypertension despite antihypertensive treatment as well as clear evidence of systemic inflammation (TNF-α). Inflammation is considered to be a main driver of cardiovascular disease in CKD^15^. Early cardiovascular pathologies and complications in children with CKD have been described previously^16,17^, the burden of which accumulates with age and causes a fatal increase of CKD-associated mortality^1^. This is remarkable given that CKD occurs in children in the virtual absence of diseases that can cause both CKD and CVD, such as diabetes and metabolic syndrome. This is particularly relevant for the present study, as traditional comorbidities in adults represent additional influences on microbiota-immune interaction. Therefore, it can be assumed that the microbiome and immune signatures presented here are influenced to a smaller degree by traditional comorbidities and represent specific signatures of CKD.

For the first time, we demonstrate that children with CKD already exhibit considerable stage-dependent intestinal barrier dysfunction indicated by increased serum levels of Zo-1 and sCD14. Zo-1 regulates intestinal permeability^18^ and has been associated with impaired barrier function in various conditions such as obesity, diabetes^19^ and autoimmune diseases^20^. Impaired barrier function permits the translocation of luminal LPS into the circulation. Consequently, we detected a CKD stage dependent increase in serum levels of the LPS binding protein sCD14, suggesting LPS-induced inflammation^21-23^. Elevated serum levels of sCD14 have been associated with an increased risk of CVD in two independent cohort studies^24,25^. Intestinal barrier dysfunction develops along with or as a consequence of dysbiosis, presumably as a consequence of a dysbalance of microbial metabolites, such as SCFA^26,27^. Therefore, we analyzed the composition of the gut microbiome in our cohort. In line with studies of adult CKD patients^5,7,28,29^, we observed CKD stage-dependent compositional changes. This microbial signature was most dominantly present in patients with HD displaying an increase of proteolytic bacteria such as *Citrobacter* (Enterobacteriaceae) and a decrease of saccharolytic bacteria such as *Bifidobacterium* (Bifidobacteriaceae), *Fusicatenibacter (*Lachnospiraceae) and *Subdoligranulum* (Ruminococcaceae). Since adults, unlike children, may well exhibit a variety of comorbidities whose presence can have an additional obscuring influence on the microbiome (e.g. diabetes^30^, obesity^31^, and fatty liver disease^32^), it can be assumed that the data presented here are less influenced by such comorbidities and therefore appear to be more clearly CKD-associated.

The enrichment of Enterobacteriaceae found in HD patients affects the microbial metabolism of nutrients as they express tryptophanases that metabolize TRP to indoles^29^. Dietary TRP is metabolized both by somatic cells and by the intestinal microbiota to metabolites with various functions^33,34^. We found that TRP metabolites in the blood of CKD patients increased in a stage-dependent manner, which was predominantly driven by the disproportional increase of metabolites of bacterial origin. Therefore, dysbiosis and intestinal barrier dysfunction, as present in CKD and HD patients in our cohort, are likely contributing to the accumulation of uremic solutes^35^ in addition to their reduced renal elimination^29,36^.

In our cohort, microbially-derived IxS and ILA as well as host-derived KYN, 3HK, KA, XA and AA were significantly elevated in CKD and HD patients, while serum levels of TRP were reduced. Both indole and KYN derivates are ligands of the AhR, thereby influencing innate and adaptive immune responses^10,33,34^. Consequently, the AhR activation potential of serum from our HD patients was increased, confirming previous findings in adults^37^. Of note, the degradation of TRP to KYN metabolites is regulated by tryptophan 2,3-dioxygenase in the liver, which is upregulated by chronic inflammation^38^ and influenced by diabetes^39-41^ and obesity^42^. In contrast to previous studies^39-41^, our study emphasizes enhanced TRP metabolism and AhR activation in CKD irrespective of traditional comorbidities.

Myeloid cells are known to be modulated by the AhR and are thus particularly affected by uremic TRP metabolites^10,33,34^. As indicated by unsupervised clustering and confirmed by hierarchical gating we demonstrated a pathological shift from classical towards intermediate and non-classical monocytes in HD patients. This monocyte signature has previously been associated with an increased risk of CVD^43,44^. Most notably, we showed that isolated monocytes exhibited a higher production of TNF-α after incubation with serum from patients treated by HD compared to HC, which was dependent on AhR function. Thus, the increased TNF-α concentrations in the circulation are at least partly a consequence of the effect of the increased and mainly microbially produced TRP metabolites on the AhR of monocytes, and possibly also other immune cells.

In line with the depletion of SCFA-generating saccharolytic bacteria^45^, such as *Bifidobacterium, Fusicatenibacter* and *Subdoligranulum* in CKD and HD patients, we show lower systemic levels of the SCFA propionate. There is a growing body of evidence about the pivotal role of SCFA in local gut homeostasis and the protection from CVD *in vitro*^*46*^ and *in vivo*^*47*^. Since SCFA are known to enhance the abundance and suppressive function of Treg cells in mice^48^ and humans^49^, we analyzed T cells by multi-color flow cytometry. Based on the expression of chemokine receptors^14,50^, we found a decrease of Th1-like, Th17-like and Th22-like Treg in HD patients. The lower frequencies of these Treg subtypes might be explained by both, lower peripheral induction as a consequence of reduced availability of SCFA^48^ and increased recruitment of Treg to sides of inflammation, e.g. atherosclerotic plaques^51^. Moreover, the frequency of dysfunctional Treg with a central memory phenotype expressing PD-1 was elevated in HD patients. Circulating Treg expressing PD-1 are known to exhibit reduced suppressive function and molecular signatures of exhaustion^52^. In light of the recognized importance of chronic inflammation for CVD development, the diminished anti-inflammatory function as a consequence of reduced abundances of Treg subtypes and increased abundances of dysfunctional, exhausted Treg is likely to play a role in CVD development^50,51^.

Interestingly, our unsupervised FlowSOM analysis clearly indicated that circulating MAIT cells were reduced in HD patients, expressed markers of exhaustion (PD-1) and produced higher amounts of IL-17A. This pattern has been described for several autoimmune, inflammatory and cardiometabolic diseases^53^, such as obesity and type two diabetes^54^. It is still a subject of discussion whether decreased MAIT cell abundances are mainly a consequence of migration to inflamed tissue or increased apoptosis^53^. In animal models, MAIT cells can promote inflammation and microbial dysbiosis leading to metabolic dysfunction during obesity^53^. A large cross-sectional analysis in patients with cardiometabolic diseases highlighted the positive association between decreased MAIT cell abundances and cardiovascular disease risk^55^. A reduction of MAIT cell abundance has been described in CKD of predominantly diabetic cause, albeit without functional cytokine expression as shown here^56^. Taken together, Treg and MAIT are two important cell populations phenotypically altered in HD patients that are known to be regulated by the microbiota, further emphasizing the importance of the microbiota-immune axis in CKD.

Noteworthy, patients after KT still showed significantly elevated serum levels of TNF-α compared to HC, albeit lower than in CKD and HD. There were no differences in serum markers of intestinal barrier function and only slight alterations in the microbiota composition, although the limited size of our cohort is less suited to detect fine-scaled differences between HC and KT. While microbial SCFA and indole metabolism apparently recovers after KT, serum levels of KYN and 3HK were still elevated. In contrast, adult patients after KT exhibited a significant decrease in alpha diversity and an increase of Proteobacteria, which is in part driven by the use of immunosuppressive drugs^57^. In a murine kidney transplantation model, mice after allograft transplantation exhibited dysbiosis even in the absence of antibiotics and immunosuppression^58^. As gut dysbiosis and impaired microbial production of SCFA seem to play a role in allograft rejection as well^58^, the absence of dysbiosis in children after KT might attribute to a more favorable transplant outcome in children compared to adults.

In conclusion, the present study is the first to show CKD stage-dependent alterations of the microbiota-metabolite-immune axis in children with CKD in the absence of comorbidities seen in adult patients. Our data demonstrate alterations at all levels of this pivotal axis. In this context, TRP metabolites act as a mechanistic link between the microbiota and the immune system, contributing to a pro-inflammatory phenotype in an AhR-dependent manner - even at a young age. SCFA deficiency may further exacerbate CKD-associated chronic inflammation and intestinal barrier dysfunction. These data provide strong evidence that the microbiota is an important stimulus for persistent inflammation. Restoring intestinal eubiosis could favorably influence the pro-inflammatory sequelae. Therefore, the microbiota appears a promising target of future therapeutic strategies aimed at sustained containment of inflammation to prevent chronic sequelae such as CVD and premature mortality in CKD.

## Data Availability

All data produced in the present study are available upon reasonable request to the authors

## Acknowledgments

This work was supported by local resources of the participating institutions. JH was supported by the Peter-Stiftung für die Nierenwissenschaft. JH, MW, SKF, and UL were supported by the German Center for Cardiovascular Research (DZHK), partner site Berlin. UL was supported by the German Federal Ministry of Education and Research (EMBARK; 01KI1909B) under the frame of JPI AMR (EMBARK; JPIAMR2019-109). S.G. was supported by the Bundesministerium für Bildung und Forschung funding MSTARS (Multimodal Clinical Mass Spectrometry to Target Treatment Resistance). F.B. was supported by the Berlin Institute of Health BIH-MD-TRENAL stipend. NW was supported by the European Research Council (ERC) under the European Union’s Horizon 2020 research and innovation program (852796) and by a grant from the Corona-Stiftung in the German Stifterverband. NW and SKF were supported by the Deutsche Forschungsgemeinschaft (German Research Foundation, SFB 1365 and SFB 1470). The sponsors had no involvement in study design, data collection, analysis and interpretation of data and in the decision to submit this article for publication.

The authors thank Gudrun Koch, Jana Czychi, Gabriele N’diaye and Kerstin Sommer for their technical assistance and Nadine Unterwalder for her kind support in analyzing serum cytokine levels.

## Author contributions

J.H., N.W. and D.M. led and conceived the project. J.H. and H.B. designed and performed most experiments, analyzed and interpreted the data. J.H., S.K., D.E. and L.D.K. conducted patient recruitment under supervision of D.M.. U.L., T.U.P.B., H.A. and S.K.F. performed the statistical analyses and data integration. J.H., H.B., M.I.W. and A.M. performed and analyzed flow cytometry. V.MP. performed most of the *in vitro* experiments. A.T. and O.D. performed 16S sequencing. D.L.V., M.K., S.G., S.K. and J.A.K. performed metabolite analysis. P.B., K.U.E., N.W., and D.M. helped with data interpretation. N.W. supervised the experiments and data analysis. J.H., H.B. and N.W. wrote the manuscript with key editing by F.B., U.Q. and K.U.E. and further input from all authors.

## Data availability

All data produced in the present study are available upon reasonable request to the authors.

