## Supplementary Information for "Inflammation in children with chronic kidney disease linked to gut dysbiosis and metabolite imbalance"

**Supplementary Figures**

**Supplementary Figure 1**: Study description

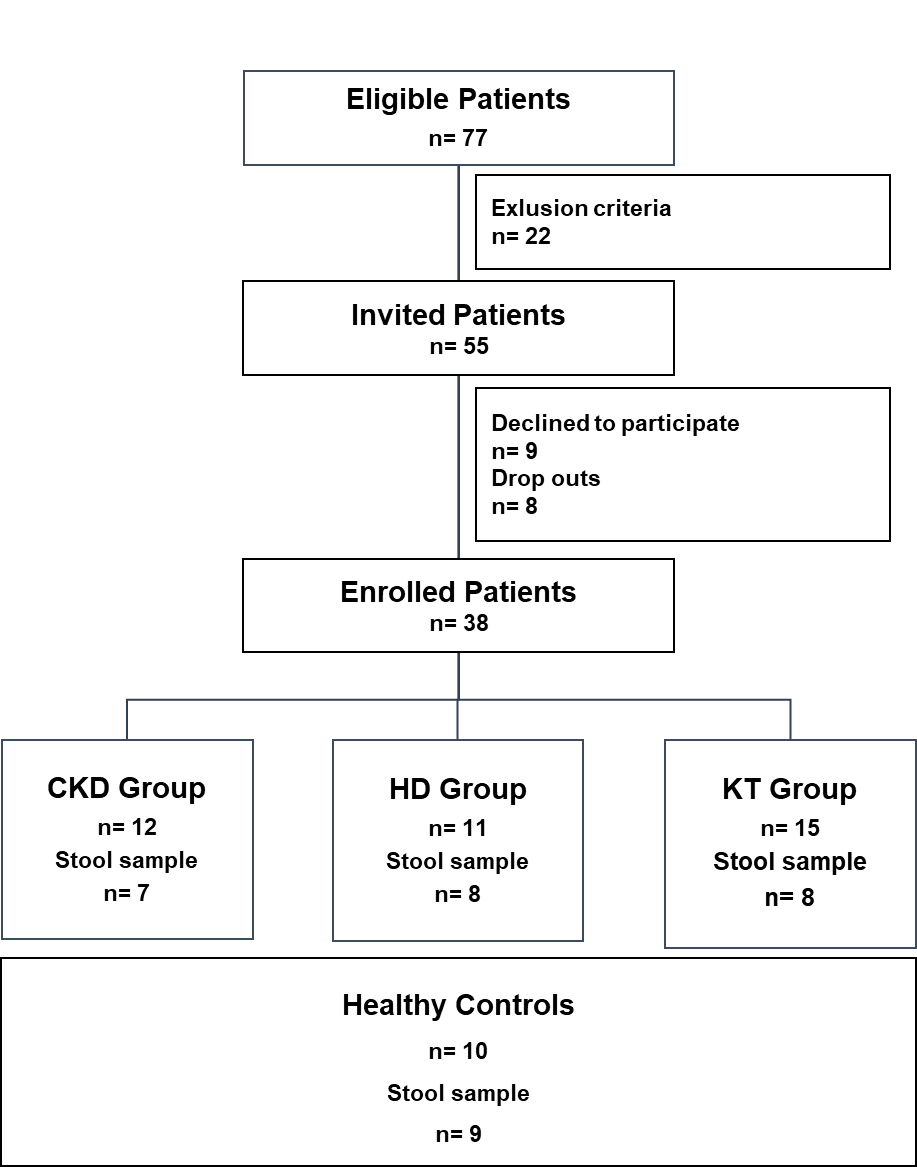

From 77 patients eligible for this study, 55 patients were invited to participate (22 were excluded as they met exclusion criteria). 9 patients or their parents declined to participate and 8 patients were excluded during the recruitment for other reasons (antibiotic treatment, lost to follow-up, difficulty in blood drawing). Moreover, 10 healthy individuals with normal kidney function, treated at the hospital for reasons other than kidney disease, were enrolled to this study.

Abbreviations: CKD = chronic kidney disease; HD = hemodialysis; KT = kidney transplantation.

**Supplementary Figure 2:** Activity of the aryl hydrocarbon receptor in response to different concentrations of indoxyl sulfate

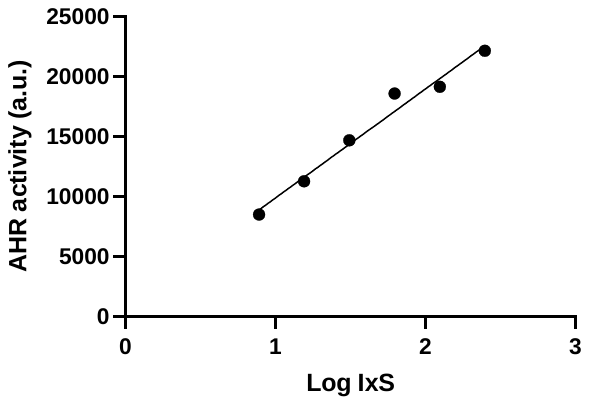

Transfected HT29 reporter cells were incubated with different concentrations of indoxyl sulfate (IxS) for 24 hours. Subsequently, the luciferase activity was measured indicating the activity of the aryl hydrocarbon receptor (AhR).

**Supplementary Figure 3**: Potential for short chain fatty acid (SCFA) production and its serum levels in children with chronic kidney disease

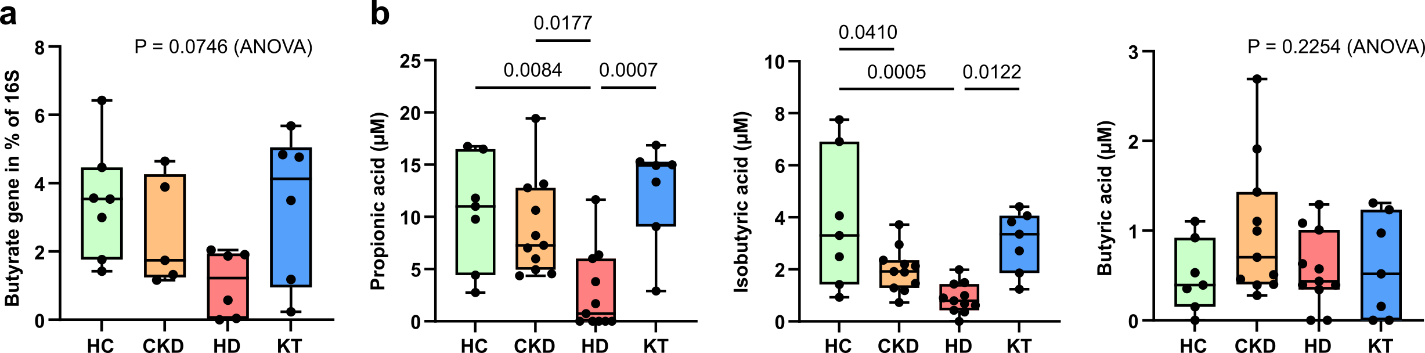
 The abundance of a butyrate associated enzymatic gene was analyzed in fecal samples (n=24) showing a lower abundance in patients of the HD group (a). Systemic levels of the SCFA propionate, isobutyrate and butyrate were analyzed in serum samples (n=36, b).  *P* values ≤ 0.05 are shown, as measured by ordinary one-way ANOVA or Kruskal-Wallis test and adjusted by post-hoc Tukey´s or Dunn´s correction for multiple testing. Data is shown as a box (median and interquartile range) and whiskers (min-max) with overlaid dot plot.

Abbreviations: CKD= chronic kidney disease, HD= hemodialysis, KT= kidney transplantation, HC= healthy controls

**Supplementary Figure 4:** Gating strategy

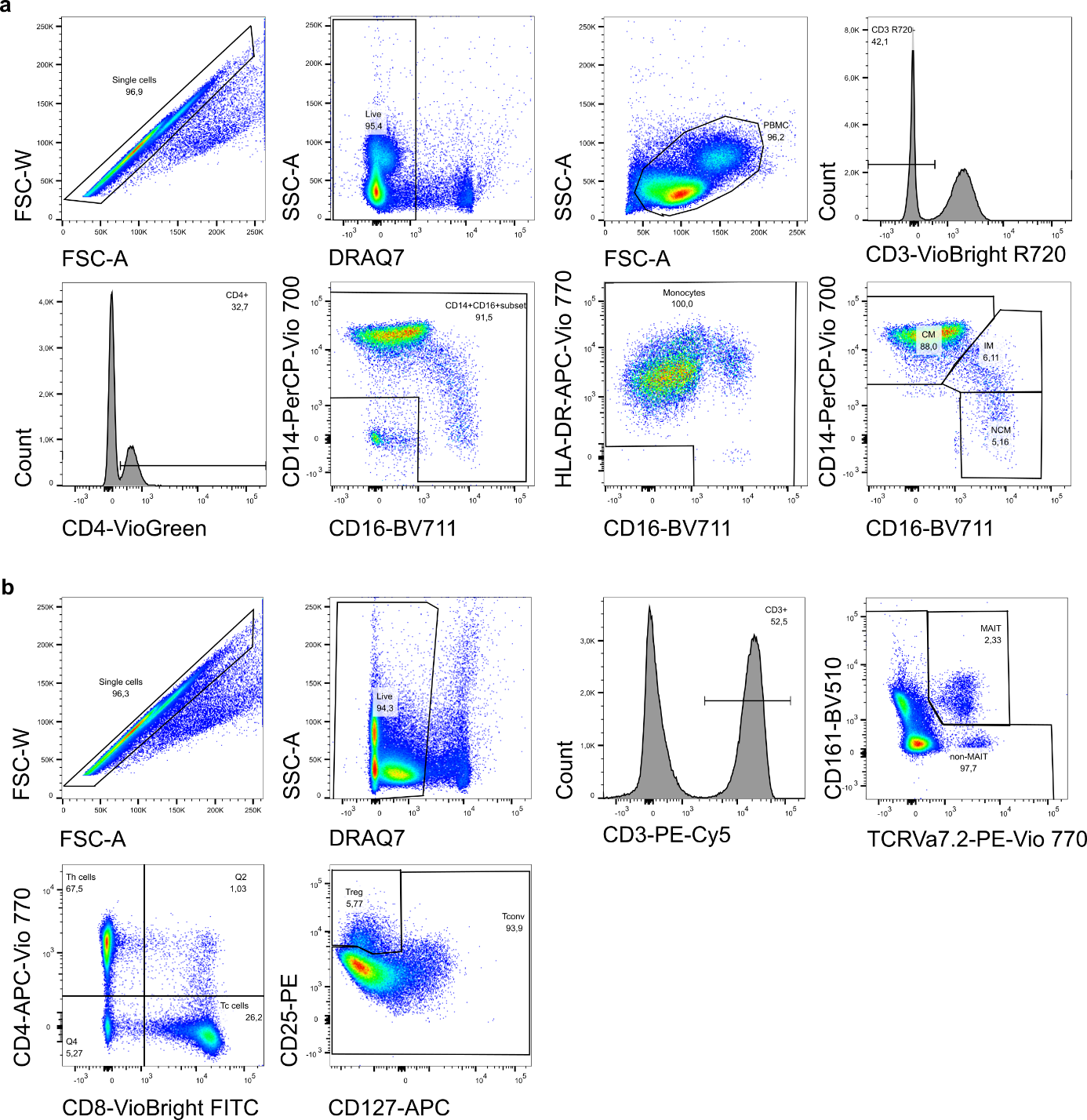

Gating strategy for the analysis of human peripheral blood mononuclear cells for flow cytometry. a) Gating for monocyte subsets. b) Gating for MAIT cells and Treg.

Abbreviations: PBMC= peripheral blood mononucleated cells; MAIT cells= mucosa associated invariant t cells; Treg= regulatory T cells; CM= classical monocytes; IM= intermediate monocytes; NCM= non-classical monocytes; Th= T helper; Tc= T cytotoxic; Tcon= T conventional.

**Supplementary Figure 5:** Dendritic cell differentiation in human peripheral blood mononuclear cells

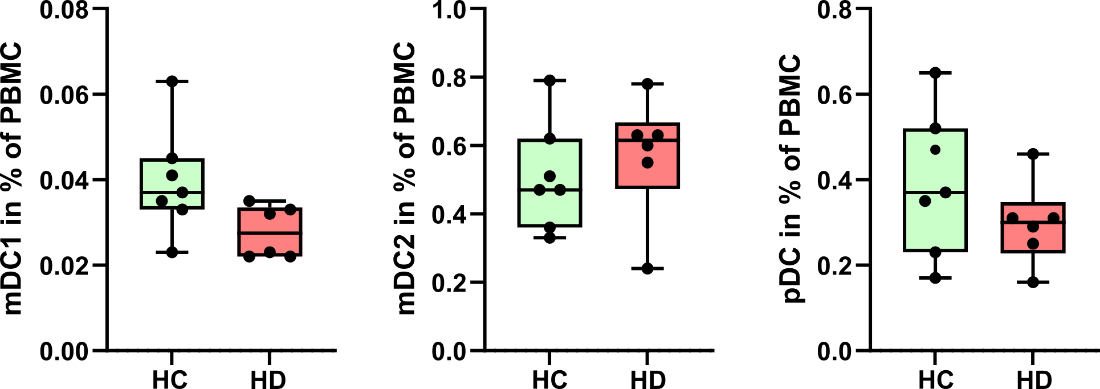

The relative abundance (in % of peripheral blood mononuclear cells (PBMC)) is shown for myeloid dendritic cells type 1 (mDC1: HLA-DR+CD141+CleC9a+), mDC2 (HLA-DR+, CD1c+, CD11c+) and plasmacytoid DC (pDC: HLA-DR+, CD123+) in HD (n=6) and HC (n=7) individuals. *P* values ≤ 0.05 are shown as analyzed by t test or Mann-Whitney-U test.

**Supplementary Figure 6:** Hierarchical gating of mucosa-associated invariant T cells and regulatory T cells

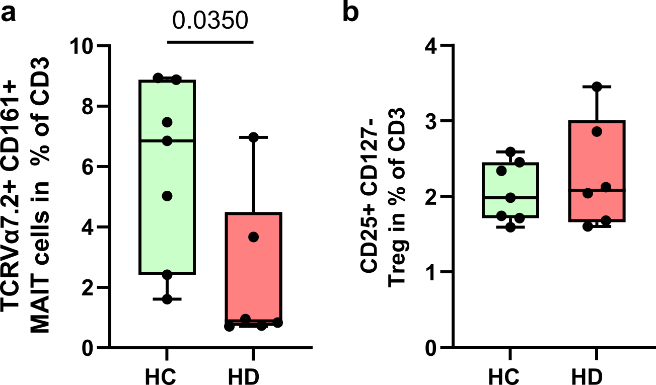

The relative abundance (in % of CD3+ T cells) is shown for mucosa-associated invariant T (MAIT) cells, defined as CD161+TCRVa7.2+ (a) and regulatory T cells (Treg), defined as CD25+CD127- in HD (n=6) and HC (n=7) individuals. *P* values ≤ 0.05 are shown as analyzed by t test or Mann-Whitney-U test.

**Supplementary Tables**

**Supplementary Table 1**: Antibodies used for flow cytometry

| **Antibodies (Monocytes/DC)** | **Dilution** | **Source** | **Identifier** |
| --- | --- | --- | --- |
| **CD123, FITC, clone REA918** | 1:50 | Miltenyi | #130-115-269 |
| **CD14, PerCP-Vio 700, clone REA599** | 1:400 | Miltenyi | #130-110-523 |
| **CleC9a, APC, clone REA976** | 1:50 | Miltenyi | #130-116-354 |
| **HLA-DR, APC-Vio 770, clone REA805** | 1:50 | Miltenyi | #130-111-792 |
| **CD3, VioBright R720, clone REA613** | 1:50 | Miltenyi | #130-127-377 |
| **CD8, BV786, clone RPA-T8** | 1:20 | BL | #301046 |
| **CD16, BV711, clone 3G8** | 1:20 | BL | #302044 |
| **CD11c, BV650, clone Bu15** | 1:20 | BL | #337283 |
| **CD141, BV605, clone M80** | 1:20 | BL | #344118 |
| **CD4, VioGreen, clone VIT4** | 1:50 | Miltenyi | #130-113-221 |
| **CD142, BV421, clone HTF-1** | 1:10 | Miltenyi | #130-098-921 |
| **CD163, PE-Vio 770, clone REA179** | 1:50 | Miltenyi | #130-112-130 |
| **DRAQ7 (L/D)** | 1:500 | Miltenyi | #130-117-342 |
| **CD1c, PE-Vio 615, clone REA694** | 1:50 | Miltenyi | #130-114-547 |
| **CD42b, PE, clone REA185** | 1:50 | Miltenyi | #130-123-725 |
| **Antibodies (T surface)** | **Dilution** | **Source** | **Identifier** |
| **CD8, VioBright FITC, clone BW135/80** | 1:50 | Miltenyi | #130-113-163 |
| **CCR10, PerCP-Vio 700, clone 1B5** | 1:25 | BD | #564772 |
| **CD127, APC, clone REA614** | 1:50 | Miltenyi | #130-113-413 |
| **CD4, APC-Vio 770, clone REA623** | 1:50 | Miltenyi | #130-113-223 |
| **CD45RA, BV786, clone HI100** | 1:50 | BD | #563870 |
| **CXCR3, BV711, clone GO25H7** | 1:20 | BL | #353732 |
| **CD27, BV650, clone L128** | 1:25 | BD | #563228 |
| **CCR6, BV605, clone GO34E3** | 1:20 | BL | #353420 |
| **CD161, BV510, clone HP-3G10** | 1:20 | BL | #339922 |
| **CCR8, BV421, clone 433-H** | 1:20 | BD | #566379 |
| **TCRVa7.2, PE-Vio770, clone REA179** | 1:10 | Miltenyi | #130-100-206 |
| **DRAQ7 (L/D)** | 1:500 | Miltenyi | #130-117-342 |
| **CD3, PE-Cy5, clone UCHT1** | 1:20 | BD | #555334 |
| **CCR4, PE-CF594, clone L291H4** | 1:20 | BL | #359420 |
| **CD25, PE, clone REA945** | 1:50 | Miltenyi | #130-115-534 |
| **Antibodies (T activation)** | **Dilution** | **Source** | **Identifier** |
| **PD-1 VioBright B515, clone REA1165** | 1:50 | Miltenyi | #130-120-386 |
| **CD8, PerCP-Vio 700, clone BW135/80** | 1:50 | Miltenyi | #130-113-161 |
| **CD69, APC, clone REA824** | 1:50 | Miltenyi | #130-112-614 |
| **CD3, APC-Vio 770, clone REA613** | 1:50 | Miltenyi | #130-113-136 |
| **BD Viability Stain 700** | 1:1000 | BD | #564997 |
| **CD45RA, BV786, clone HI100** | 1:25 | BD | #563870 |
| **CD31, BV711, clone WM59** | 1:25 | BL | #303136 |
| **CD161, BV650, clone DX12** | 1:20 | BD | #563864 |
| **CD39, BV605, clone A1** | 1:25 | BL | #328236 |
| **CD25, BV510, clone M-A251** | 1:20 | BD | #563352 |
| **TCRVa7.2, VioBlue, clone REA179** | 1:10 | Miltenyi | #130-100-209 |
| **CD127, PE-Vio 770, clone REA614** | 1:50 | Miltenyi | #130-113-415 |
| **CD4, PE-Cy5, clone RPA-T4** | 1:25 | BD | #555348 |
| **CD62L, PE-CF594, clone DREG-56** | 1:25 | BD | #562301 |
| **FoxP3, PE, clone 259D** | 1:20 | BL | #320108 |
| **Antibodies (MAIT cytokines)** | **Dilution** | **Source** | **Identifier** |
| **TCRVa7.2 FITC, clone REA179** | 1:50 | Miltenyi | #130-123-685 |
| **IL22, APC, clone REA466** | 1:10 | Miltenyi | #130-111-768 |
| **IL17A, APC-Vio 770, clone CZ8-23G1** | 1:50 | Miltenyi | #130-120-409 |
| **TNFa, R718, clone Mab11** | 1:1:20 | BD | #566957 |
| **CD3, BV786, clone SK7** | 1:100 | BD | #563870 |
| **IFNg, BV711, clone B27** | 1:20 | BD | #564039 |
| **CD8, BV650, clone SK1** | 1:100 | BL | #344730 |
| **IL2, BV605, clone 5344,111** | 1:20 | BD | #563947 |
| **Ki-67, BV510, clone B56** | 1:20 | BD | #563462 |
| **GRZB, BV421, clone GB11** | 1:400 | BD | #563389 |
| **CD161, PE-Vio 770, clone REA631** | 1:50 | Miltenyi | #130-113-597 |
| **CD4, PE-Cy5, clone RPA-T4** | 1:20 | BD | #555348 |
| **BD Viabilty Stain 620** | 1:500 | BD | #564996 |
| **GMCSF, PE, clone REA1215** | 1:50 | Miltenyi | #113-123-419 |

**Supplementary Table 2**: Markers of inflammation in children with CKD and healthy individuals at study enrolment

|  | **CKD G3-4** | **HD** | **KT** | **HC** |
| --- | --- | --- | --- | --- |
| **Patients (n)** | 12 | 11 | 14 | 9 |
| **TNF-a** | 7.54 ± 2.22 | 9.63 ± 2.73 | 5.70 ± 1.66 | 2.87 ± 1.10 |
| **Interleukin 2** | 0.53 ± 0.68 | 0.34 ± 0.0 | 0.42 ± 0.32 | 0.40 ±0.17 |
| **Interleukin 4** | 0.05 ± 0.002 | 0.06 ± 0.01 | 0.05 ± 0 | 0.06 ± 0.01 |
| **Interleukin 6** | 1.06 ± 0.84 | 1.39 ± 1.03 | 1.05 ± 0.94 | 0.73 ± 0.65 |
| **Interleukin 8** | 6.52 ± 3.18 | 7.21 ± 3.58 | 10.63 ± 10.81 | 5.39 ± 1.85 |
| **Interleukin 10** | 0.54 ± 0.49 | 0.44 ± 0.33 | 0.72 ± 0.76 | 0.60 ± 0.48 |
| **Interleukin 13** | 0.19 ± 0.15 | 0.18 ± 0.10 | 0.35 ± 0.40 | 0.30 ± 0.34 |
| **Interleukin 1-β** | 0.19 ± 0.18 | 1.00 ± 2.13 | 0.16 ± 0.08 | 0.59 ± 1.35 |
| **Interleukin 12p70** | 0.22 ± 0.15 | 0.19 ± 0.11 | 0.21 ± 0.16 | 0.18 ± 0.07 |
| **Interferon y** | 1.06 ± 0.84 | 1.27 ± 1.06 | 5.05 ± 7.28 | 6.37 ± 10.2 |
| **CrP** | 0.79 ± 1.03 | 1.71 ± 1.60 | 2.01 ± 1.81 | 2.64 ± 5.93 |

Cytokines were measured in 46 patients at time of study enrolment. Patients were grouped into four categories (CKD= chronic kidney disease, HD= hemodialysis, KT= kidney transplantation, HC= healthy controls). Data is shown as mean ± standard deviation.

**Supplementary Table 3**: Multivariable ANOVAs on possible confounders influencing the serum levels of tryptophan metabolites in children with chronic kidney disease

| **Metabolite** | **Age**  **(years)** | **BMI**  **(kg/m^2^)** | **Disease** | **eGFR**  **(ml/min/1.73m^2^)** | **Group** | **Ethnic Bachground** | **Gender** |
| --- | --- | --- | --- | --- | --- | --- | --- |
| **3OH-Kynurenin** | 0.56 | 1 | 1 | 0.23 | <0.001 | 0.23 | 1 |
| **5OH-Tryptophan** | 1 | 1 | 1 | 0.02 | 0.72 | 1 | 0.44 |
| **Anthranilic acid** | 1 | 1 | 1 | 1 | <0.001 | 1 | 1 |
| **Indole-3-carboxyaldehyde** | 1 | 1 | 1 | 0.99 | 1 | 1 | 1 |
| **Indole-3-propionate** | 1 | 1 | 1 | 1 | 1 | 1 | 1 |
| **Indole lactate** | 1 | 1 | 1 | 0.06 | <0.001 | 1 | 1 |
| **Indoxyl sulfate** | 1 | 1 | 1 | 1 | <0.001 | 1 | 1 |
| **Kynurenin/Tryptophan** | 1 | 1 | 1 | 1 | <0.001 | 1 | 1 |
| **Kynurenic acid** | 1 | 1 | 1 | 1 | <0.001 | 1 | 1 |
| **Kynurenine** | 1 | 1 | 1 | 1 | 0.001 | 1 | 1 |
| **Tryptamin** | 1 | 1 | 1 | 1 | 1 | 1 | 1 |
| **Tryptophan** | 1 | 1 | 1 | 1 | <0.001 | 1 | 1 |
| **Xanthurenic acid** | 1 | 1 | 1 | 1 | 0.001 | 0.61 | 1 |

Multivariable ANOVAs revealed a significant influence of patient group (CKD, HD, KT, HC) on the plasma levels of tryptophan metabolites in 48 children with CKD. Age, BMI, underlying kidney disease, eGFR, ethnic background and gender had no influence on plasma metabolite levels. All significance estimates were adjusted for multiple tests using Benjamini-Hochberg FDR correction.

Abbreviations: BMI= body mass index, eGFR= estimated glomerular filtration rate, CKD= chronic kidney disease, HD= hemodialyis, KT= kidney transplantation, HC= healthy controls.

**Supplementary Methods**

**Clinical assessment, biobanking and routine laboratory measurements**

At the time of enrolment, we obtained baseline demographic (age, gender, diagnosis, ethnic background, body weight and height) and clinical data from all patients. eGFR was calculated according to the bedside formula of Schwartz based on serum creatinine^1^, percentiles of weight and BMI were determined according to national references^2^. Office systolic and diastolic blood pressures were documented as an average of three oscillometric measurements using local devices and normalized to national references^3^. Arterial hypertension was defined as blood pressure values above the 95^th^ percentile.

Heparinized blood specimens were collected as part of routine laboratory sampling and used for the measurement of creatinine, urea, uric acid, phosphate, albumin, C-reactive protein (CrP), parathyroid hormone (PTH), and triglyceride levels. Serum, EDTA and Heparin plasma were stored at -80°C until further use. For the measurement of TNF-α, IL-2, IL-4, IL-6, IL-8, IL-1β, IL-10, IL-13, IL-12p70, and IFN-y, plasma was analyzed using the Meso Scale Discovery (MSD) V-PLEX Plus Proinflammatory Panel 1 (human) according to the manufacturer’s protocol. All samples were run on a single plate on an MSD plate reader (model 1250). Zonulin-1 (Zo-1) and soluble CD14 (sCD14) were analyzed in patients serum using the human Zo-1 ELISA kit (Biomatik Corporation, Canada) and sCD14 Quantikine ELISA kit (R&D Systems, USA) according to manufacturer’s protocol.

Peripheral blood mononuclear cells (PBMC) were isolated by density gradation using Pancoll (Pan Biotech, Germany) using standard protocols and stored in liquid nitrogen until usage.

Stool specimens (2-4g) were collected (Sarstedt, Germany; #80.623.022) and stored for 24 hours at 4 - 8°C max. and transferred to the study center for freezing at -80°C until further use. Patients and parents were provided with detailed information about collection, storage and transport of stool specimens.

**Generation of 16S rRNA Amplicon Libraries and Sequencing**

Stool DNA was isolated using QIAamp® Fast DNA Stool Mini Kit (QIAGEN, Hilden, Germany) according to manufacturer’s protocol. The extracted DNA was used as a template to amplify the V3-V4 region of the bacterial 16S rRNA gene using fusion primers TCGTCGGCAGCGTCAGATGTGTATAAGAGACAG-CCTACGGGNGGCWGCAG (MiSeq_overhang-D-Bact-0341-b-S-17) and GTCTCGTGGGCTCGGAGATGTGTATAAGAGACAG-GACTACHVGGGTATCTAATCC (MiSeq_overhang-S-D-Bact-0785-a-A-21) including bacteria targeting primers^4^. The PCR reaction mixture with a total volume of 25 µl contained 12,5 µl KAPA HiFi HotStart ReadyMix (Roche), 1,25 µl of each primer (10 nM) and 3 ng/µl of isolated DNA. Thermal cycling scheme for bacterial amplicons was as follows: initial denaturation for 3 min at 95°C, 25 cycles at 95°C for 30 s, 30 s at 55°C, and 30 s at 72°C and a final extension at 72°C for 5 min. For each sample, three independent 16S amplicons were generated. The resulting PCR products were purified with Agencourt AMPure XP magnetic beads (Beckman coulter, Krefeld, Germany) and then quantified with Quant-iT dsDNA HS assay kit and a Qubit fluorometer (Invitrogen GmbH, Karlsruhe, Germany) following the manufacturer´s instructions. The three individual, purified V3-V4 PCR products per sample were pooled and used to attach indices and Illumina sequencing adapters using the Nextera XT Index kit (Illumina, CA, USA). Index PCR was performed using 5 μl of template PCR product (each 2 ng/µl), 2.5 μl of each index primer, 12.5 μl of 2× KAPA HiFi HotStart ReadyMix and 2.5 μl PCR grade water. Thermal cycling scheme was as follows: 95°C for 3 min, 8 cycles of 30 s at 95°C, 30 s at 55°C and 30 s at 72°C, and a final extension at 72°C for 5 min. Bacterial 16S Amplicon libraries were sequenced using the dual index paired-end (v3, 2×300 bp) approach for the Illumina MiSeq platform as recommended by the manufacturer.16S amplicon sequences were filtered, quality controlled and taxonomically assigned using the LotuS pipeline^5^ with default parameters using the SILVA database (v138). Resulting abundance tables were normalized using the rarefaction toolkit (RTK) using default settings^6^.

**Real Time PCR**

PCRs were carried out using a Quantstudio 3 (Applied Biosciences, CA, USA) thermal cycler in a reaction volume of 20µl Taqman fast universal master mix (Applied Biosciences) with Taqman gene expression assay for butyrate associated bacterial genes. 50ng of bacterial genomic DNA was used as template and all qPCR was carried out at 60°C in fast mode and primer and probe concentrations were set to 750nM and 250nM, respectively. Gene abundancy was determined relative to bacterial 16s rRNA gene.

**Targeted metabolomics**

The analysis of plasma metabolites was focused on tryptophan metabolites and short chain fatty acids (SCFA). For the tryptophan analysis, liquid chromatography – mass spectrometry (LC-MS) analysis was performed with a 1290 Infinity 2D HPLC system (Agilent Technologies, USA) combined with a TSQ Quantiva triple quadrupole mass spectrometer with a heated ESI source (Thermo Scientific, USA). Before starting, an extracting solvent was prepared comprising 90% methanol, 0.15 µg/mL mixed internal standards, 0.02% ascorbic acid, and 0.2% formic acid. This was placed at -20°C to cool. For each sample, 280 µL pre-chilled extracting solvent was added to 150ul of EDTA plasma. Samples were held at 4°C and shaken for 15 min at 2000rpm (Eppendorf ThermoMixer C) before being centrifuged for 15 min at 11000g and 4 ^o^C. The supernatant was transferred to a dark LC-MS vial for LC-MS/MS analysis. 20µl of each plasma sample was pooled, and the pooled plasma was also extracted to make quality control (QC) samples. These QC samples were run every 6 samples.

LC-MS analysis was combined with a triple quadrupole mass spectrometer using a 10-min gradient. A reversed-phase column was used (VisionHT C18 Basic; L × I.D. 150 mm × 4.6 mm, 3 μm particle size) and held at a constant temperature of 30°C. The mobile phase consisted of 0.2 % formic acid in H_2_O (solvent A) and 0.2 % formic acid in methanol (solvent B). The following gradient was run with a constant flow rate of 0.4 ml min^−1^: A/B 97/3 (0 min), 70/30 (from 1.2 min), 40/60 (from 2.7 to 3.75 min), 5/95 (from 4.5 to 6.6 min) and 97/3 (from 6.75 to 10 min). Before starting, the LC-MS was conditioned by running 10 blank (solvent A) and 3 QC samples for 130 min. The molecular ion and at least two transitions were monitored for up to 32 metabolites that comprise the tryptophan pathway.

Data was exported into Skyline v.19.1 - 64 bit to identify and quantify peak intensity and area. Transition settings in the Skyline search were: isotopic peaks included: count; precursor mass analyzer: QIT; acquisition method: targeted; product mass analyzer: QIT. Method match searching tolerance was 0.6 m/z, and data was manually checked to ensure the correct peaks were selected. Cubic spline drift correction was applied per metabolite and to all sample types using the pooled QC samples as references to fit the splines^7, 8^. The first and the last QC samples used to fit the cubic splines are the most critical to the resulting fit. In this case, the last conditioning pooled QC sample and the first of two pooled QC replicates at the end of the analytical run were used as the first and last QC respectively in the batch correction. QCs analyzed before or after these timepoints were disregarded for further analysis. For indole-lactate, both end replicates had to be removed as outliers due to clear deviation in areas, and further processing was limited to samples up to the last valid pooled QC sample. Standard samples of increasing concentration were used to construct calibrations curves using linear fits per metabolite. Concentration values less or equal to zero were declared as missing. In this study, several study groups featured measurements systematically outside of the calibration range for some metabolites (i.e. below or above the smallest or largest standard sample applied for a metabolites calibration curve, respectively). Further, calibration of QC standard samples resulted in insufficient accuracy. However, precision (%RSD in either standard or pooled QC samples) was adequate for most compounds. Hence, metabolites can not be considered as absolutely but as relatively quantified in this study. Three out of 15 TRP metabolites (tryptophan, 5-OH-tryptophan, tryptamin, kynurenine, 3-OH-kynurenine, kynurenic acid, anthranilic acid, xanthurenic acid, IxS, indole-3-carboxyaldehyde, indole-3-propionic acid, indole lactate, 6-OH-melatonin, melatonin, serotonin) were rejected due to relative standard deviation (RSD) higher than 15%, including 6-OH-melatonin, melatonin and serotonin.

SCFA were analyzed by gas chromatography (GC – MS). In brief, 90µl serum were mixed with internal standard (100µM crotonic acid (Sigma Aldrich)), 10µl HCL-37% (Sigma Aldrich) and 100 μL diethylether (Sigma Aldrich). Samples were shaken for 30 min at 1500 rpm at 25°C and then centrifuged for 10 min at 1500×g. 50µl of the upper organic phase were transferred to a Chromacol^TM^ glass vial (Thermo Scientific), containing 10µl N-tert-butyldimethylsilyl-N-methyltrifluoroacetamide (MTBSTFA, Sigma Aldrich). Samples were shaken for 30 min at 80°C at 600rpm and subsequently incubated at RT for 24 hours for derivatization. Serial dilutions of SCFA were processed and analysed in parallel for absolute quantification.

GC-MS analysis was performed on a Trace 1310 GC – Q Exactive MS, coupled to a TriPlus RSH autosampler (Thermo Fisher Scientific). Samples were injected in split mode (injection volume 1µL, split 1:10). The following temperature program was applied during sample injection: initial temperature of 80°C for 3 s followed by a ramp of 7°C/sec to 210°C and final hold for 3 min. Gas chromatographic separation was performed with a TG-5SILMS column (30m length, 250µm inner diameter, 0.25µm film thickness (Thermo Fisher)). Helium was used as carrier gas with a flow rate of 1.2 ml/min. Gas chromatographic separation was performed with the following temperature gradient: 2 min initial hold at 68°C, first temperature gradient with 7°C/min up to 150°C, and second temperature gradient with 50°C/min up to 300°C, with a final hold for 2 min. The spectra were recorded in a mass range of 65 to 600m/z with a resolution of 60,000.

Data was analyzed with Xcalibur QuantBrowser, using serial dilutions of SCFA (ranging from 500nM to 50µM) as well as internal standard (crotonic acid) calibration for quantification (linear fit of area-ratio (SCFA/InternalStandard) to concentration, with 1/X weighting). Following masses were quantified: 131.0524 m/z for propionate, 145.0680 m/z for iso-butyrate and butyrate, and 143.0523 m/z for crotonic acid.

**Aryl hydrocarbon receptor (AhR) activity assay**

HT-29-AhR cell line (InvivoGen, CA, USA) was grown and maintained in Dulbecco’s Modified Eagle Medium (DMEM, Sigma Aldrich) supplemented with 4.5g/l glucose (Sigma Aldrich), 2mM L-glutamine (Sigma Aldrich), 10% (v/v) heat-inactivated fetal bovine serum (Biochrom, Germany), 100 µg/ml Zeocin (Sigma Aldrich) and 100 ug/ml Penicillin-Streptomycin (Sigma Aldrich). For the AhR reporter assay cells at 80% confluency were harvested and re-suspended at 2.8 x 10^5 cells/ml. 20µl of patient serum was mixed with 180µl of cell suspension in a white 96 well plate (Corning, NY, USA). The plate was incubated at 37°C with 5% CO2 for 48h. After 48h, 20µl of cell supernatant was taken from each well and used in a Quanti-Luc assay according to manufacturer’s instructions (InvivoGen). Assays were performed in triplicates and luciferase activity was quantified as relative luminescence units (RLU) using a microplate reader (Infinite 200 plate reader, Tecan, Switzerland). The AhR activity was normalized to non-inoculated media.

**Monocyte isolation and serum incubation**

PBMC were isolated from a healthy donors and monocytes were enriched using CD14 MicroBeads (Milteny Biotec, USA) according to manufacturer´s protocol. On a 96 well plate 1x10^6^ cells were plated and incubated for 24 hours at 37°C in standard culture medium containing 20% patient serum, IxS (IxS potassium salt, Sigma Aldrich, USA) at concentrations of 50µM and 125µM or the synthetic AhR antagonist (CH-223191, Sigma Aldrich, USA) respectively. Afterwards, TNF-α was measured in the supernatant using a human TNF-α ELISA kit (Thermo Fisher, USA) according to manufacturer´s protocol.

**Flow cytometry analysis**

PBMC were analysed by multi-colour flow cytometry. To avoid batch effects, all collected cells were thawed and measured at the same time. After thawing, cells were either stained immediately in FACS buffer or restimulated in a final volume of 200 μl RPMI 1640 (Sigma-Aldrich, USA)) supplemented with 10% FBS (Merck), 100 U/ml penicillin (Sigma-Aldrich, USA), 100 mg/ml streptomycin (Sigma-Aldrich, USA), 50 ng/ml PMA (Sigma-Aldrich, USA), 250 ng/ml ionomycin (Sigma-Aldrich, USA) and 1.3 μl/ml Golgistop (BD, USA). Cells were fixed and permeabilized (eBioscience™ Foxp3/Transcription Factor Staining Buffer Set, Thermo Fisher, USA) and labelled using monoclonal antibodies (Supplemental Table 1). Cells were analysed using a LSRFortessa flow cytometer and FACSDiva software (BD, USA). Data analysis was performed with FlowJo (LLC, USA).

For FlowSOM analysis cells were manually gated (live single cells) on T cells (CD3+) and monocytic cells (CD3- CD4+) and exported for the respective analysis. This resulted in 1.1 - 6.2 x10^5^ monocytic cells per sample and 1.4 - 6.6 x10^5^ T cells per sample for further analysis. Markers used for FlowSOM analysis of the respective panel are shown in the heatmap (Figure 5c and 6b). Data were clustered onto a 10 node × 10 node square SOM as implemented in the CATALYST package v.1.14.1 (wrapper for FlowSOM). Lower resolution ConsensusClusterPlus metaclustering was then performed to reduce them to 8 metaclusters. Phenotypic relationship between individual clusters were explored using the UMAP algorithm for dimension reduction of 10^5^ gated cells from all samples acquired, as implemented in the runDR, runUMAP functions from the CATALYST package (R version 4.0.3). Resource intensive computation has been performed on the HPC for Research cluster of the Berlin Institute of Health.

**References to the Supplementary Material**
